# Pulsed Electromagnetic Fields in Chronic Pain Management: a KDM6B-Mediated Modulation Mechanism Hypothesis

**DOI:** 10.64898/2026.03.23.26348899

**Authors:** Angelo Ferraro, Cecilia Sacco

## Abstract

**Background:** Chronic pain affects millions of patients globally and remains therapeutically challenging. While conventional pharmacological approaches have limitations and side effects, pulsed electromagnetic field (PEMF) therapy represents a non-invasive biophysical approach. However, the biological mechanisms underlying PEMF efficacy remain poorly understood.

**Objective:** This study starting from a multi-center post-market surveillance (PMS) data of 81 patients treated with SynthéXer (a CE-marked Class IIa PEMF device) proposes a mechanistic framework that links observed clinical effects to epigenetic modulation via the histone demethylase KDM6B.

**Materials and Methods:** Patients with inflammatory and degenerative disorders causing chronic pain were treated with SynthéXer across four Italian rehabilitation centers. Pain was assessed using the Numerical Pain Rating Scale (NPRS) before and after treatment. Statistical analysis included descriptive statistics, ANOVA, correlations, and Cohen’s d effect size. Proposed mechanisms were based on and extrapolated from molecular and biochemical studies demonstrating KDM6B-dependent epigenetic changes in response to specific PEMF sequences.

**Results:** Mean NPRS score decreased significantly from 8.07 ± 1.65 (PRE) to 1.79 ± 1.67 (POST), representing a 6.28-point reduction (p < 0.001; Cohen’s d = 3.1). Ninety-eight percent of patients showed pain reduction ≥2 points. No adverse effects were reported. Subset analysis revealed consistent responses across inflammatory (n=19) and degenerative (n=62) pathologies.

**Discussion:** While the observational nature of these data precludes definitive causal attribution, the magnitude of clinical response combined with emerging evidence of KDM6B-mediated epigenetic remodeling suggests a plausible biological basis for PEMF efficacy. Specifically, sequence-dependent electromagnetic stimulation may promote the production of and release of anti-inflammatory cytokines and pain resolution through histone demethylation and chromatin remodeling ultimately acting on the expression modulation of such regulatory cytokines.

**Conclusions:** These post-market surveillance data provide clinical evidence of PEMF effects in chronic pain management. The proposed epigenetic mechanism, while requiring further experimental validation and mechanistic confirmation, offers a science-based framework for understanding PEMF biological action and guiding future investigations.

**Trial Registration:** This is part of a retrospective observational post-market surveillance study conducted in compliance with EU MDR 2017/745 requirements.

## 1. Introduction

Chronic pain is a complex, multifactorial condition affecting an estimated 20% of the adult population worldwide and representing a leading cause of disability and reduced quality of life [1,2]. Beyond its personal impact, chronic pain imposes substantial economic burden on healthcare systems and society [3,4]. While pharmacological interventions, particularly opioids and non-steroidal anti-inflammatory drugs (NSAIDs), remain standard of care, their efficacy is often limited, and long-term use is associated with significant adverse effects including opioid dependency, gastrointestinal complications, and cardiovascular risks [5,6]. These therapeutic gaps motivated investigation of complementary non-pharmacological, non-invasive approaches.

Pulsed electromagnetic field (PEMF) therapy is a non-thermal, non-ionizing biophysical modality that has been employed clinically for several decades, with FDA approval for bone fracture treatment dating back to 1970s following pioneering work by Bassett et al. [7]. Over the past two decades, accumulating clinical evidence and mechanistic research have expanded potential applications of PEMF to inflammatory and degenerative conditions affecting joints, soft tissues, and peripheral nerves [8,9].

The biological effects of PEMF appear to be highly dependent on electromagnetic signal characteristics—including frequency, waveform, modulation pattern, amplitude, and temporal patterning [10]. This sequence-specificity suggests that PEMF acts through structured biochemical and biophysical mechanisms rather than through non-specific thermal effects [11,12].

### 1.1 Proposed Mechanisms of PEMF Action

Several theoretical models have been proposed to explain PEMF–cell interactions. Early biophysical models emphasized ion resonance, Lorentz force effects on mobile ions, and perturbation of electrochemical gradients at the plasma membrane [11, 13–15]. More recent frameworks suggest that PEMF-induced membrane perturbations trigger signal transduction cascades affecting intracellular calcium dynamics, reactive oxygen species (ROS) production, and gene expression [16,17].

Accumulating evidence indicates that PEMF modulates expression of inflammatory mediators and cytokines in immune cells, particularly in mononuclear cells and macrophages [18– 21]. These changes include altered expression of pro-inflammatory cytokines (TNF-α, IL-1β, IL-6) and upregulation of anti-inflammatory mediators (IL-4, IL-10) [22–25]. However, the upstream molecular events linking PEMF exposure to these transcriptional changes remain partially elucidated.

### 1.2 Epigenetic Regulation and Immune Function

Recent investigations have identified epigenetic mechanisms—particularly histone modifications and chromatin remodeling—as critical nodes in the regulation of macrophage polarization and immune homeostasis [26–28]. Among key epigenetic regulators, histone demethylases of the KDM family have emerged as important modulators of immune cell differentiation and inflammatory responses [29,30].

KDM6B (also known as JMJD3) specifically catalyzes removal of repressive tri-methylation marks from histone H3 lysine 27 (H3K27me3), thereby opening chromatin structure and facilitating transcription of specific gene sets [31]. Notably, KDM6B activity has been linked to macrophage M2 (antiinflammatory) polarization and expression of anti-inflammatory interleukins [32–35]. Given the role of dysfunctional macrophage polarization in chronic inflammatory pain conditions [36], KDM6B represents a plausible molecular target through which PEMF might exert therapeutic effects.

### 1.3 Rationale for the Current Study

Recent in vitro investigations have demonstrated that exposure of human monocytic cells to specific PEMF sequences generated by the SynthéXer system induces KDM6B upregulation, H3K27 demethylation, macrophage differentiation toward an anti-inflammatory phenotype, and increased secretion of anti-inflammatory interleukins [37,38]. These findings, coupled with decades of clinical observations of PEMF efficacy in pain management, suggest an opportunity to link post-market surveillance data with emerging molecular mechanisms.

The present study accomplishes three objectives: (1) to present multi-center post-market surveillance (PMS) data documenting clinical response to PEMF therapy in a heterogeneous cohort of chronic pain patients; (2) to propose a mechanistic framework linking observed clinical effects to PEMF sequence-dependent epigenetic modulation; and (3) to identify key questions requiring further investigation to validate and extend this framework.

## 2. Materials and Methods

### 2.1 Study Design and Regulatory Context

This retrospective observational PMS study was conducted in compliance with the European Union Medical Device Regulation (MDR) 2017/745/EU. It was designed to fulfil post-market surveillance (PMS) obligations for the SynthéXer system, a CE-marked Class IIa medical device (PN E01S01025-02) manufactured by Ethidea S.r.l. (Mathi, Turin, Italy). The study consists of a retrospective observational analysis of fully anonymized PMS data collected from private centers during routine clinical use of the CE-marked device, without any additional procedures or deviations from standard practice. The manufacturer received only fully anonymized data; no direct or indirect identifiers were accessed, and no re-identification key was available to the investigators. As the dataset was fully anonymized and collected for regulatory PMS purposes, ethics committee approval and informed consent were not required, in accordance with applicable institutional and local policies governing analyses of anonymized surveillance and quality data.

### 2.2 The SynthéXer® System

SynthéXer® E01S01025-01 (Ethidea Srl, Turin, Italy) is a medical device in Class IIa, able to generate and deliver programmable ultra-stable low-intensity electromagnetic signals (0.001-2 Gauss) in the frequency range from 10 Hz to 10 kHz. The device comprises a Generator unit and an Applicator (Figure 1). The Generator is a programmable arbitrary waveform synthesizer with a built-in amplified output stage. Its intended use is PEMF therapy for the treatment of bone fractures healing and osteoarticular problems in general, osteoporosis and tendinopathy, inflammation of nerves and tissues, and pain affecting the musculoskeletal system. The Applicator is a flexible coil antenna integrated into the treatment bed, available in different geometric configurations to optimize electromagnetic field uniformity and distribution over the full body space.

**Fig. 1.**
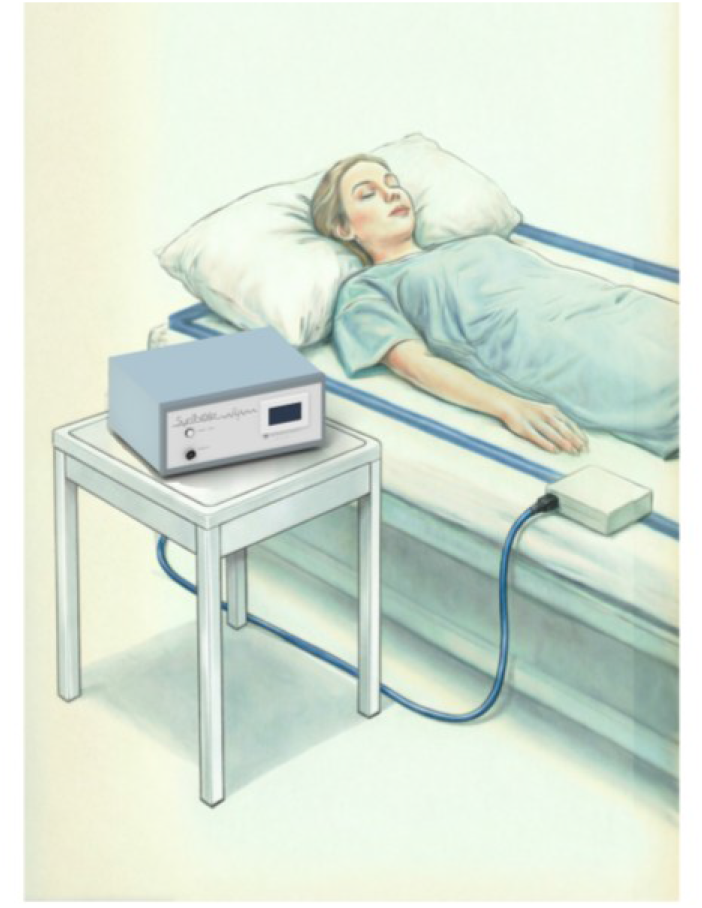
The Generator is connected to the flexible antenna (Applicator) laying on the bed, around the patient.

The distinctive feature of SynthéXer is its ability to generate complex, time-patterned sequences with precisely controlled waveform parameters (frequency, duty cycle, burst pattern, modulation envelope) (Figure 2). This capability enables delivery of “dose-field” sequences with reproducible characteristics, allowing investigation of sequence-specific biological effects. Pre-configured treatment protocols specific to different pathologies are selected by clinical staff, minimizing inter-operator variability and ensuring protocol fidelity.

**Fig. 2.**
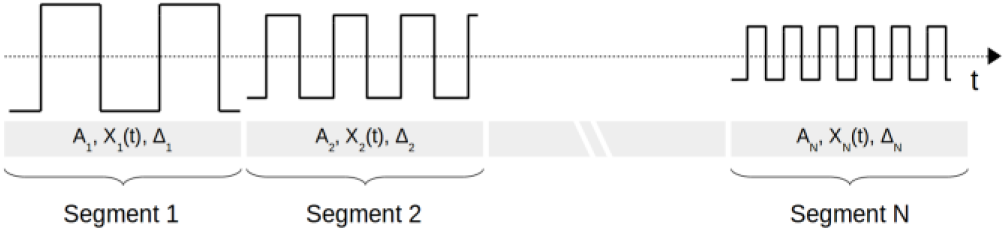
The general frame of SynthéXer^®^ signal is based on a series of N wave segments, where every segment holds a specific duration time. Within each segment several technical parameters such as frequency, waveform X_i_, modulation and amplitude A_i_ are defined.

### 2.3 Patients cohorts

This analysis includes 81 patients with chronic pain from four Italian medical centers: Arezzo (AR), Milazzo (ME), Turin (TO), Somma Lombardo (VA). **Inclusion Criteria:** Patients presenting chronic pain due to one or more of the following conditions: a) Osteo-articular disorders: fractures, osteoarthritis, rheumatoid arthritis, osteopathies, chondropathies, discopathies, osteoporosis, cervicalgia, musculo-tendinous disorders b) Rheumatological disorders c) Soft tissue disorders: gastro-abdominal-pelvic dysfunction, recurrent colitis, Crohn’s disease d) Asthma e) Food intolerances. **Exclusion Criteria:** Patients with pacemakers or other active implantable medical devices.

#### Data Processing

To ensure group homogeneity and comparability, patients with asthma and food intolerances (n=2) were excluded from final analysis. Remaining patients (n=81) were then clinically stratified into two subgroups by physician assessment (Figure 3): Inflammatory group (Phlogistic): n=19, characterized by inflammatory/soft tissue pathologies and rheumatological disorders and Degenerative group (Degenerative): n=62, characterized by osteo-articular disorders.

**Fig. 3.**
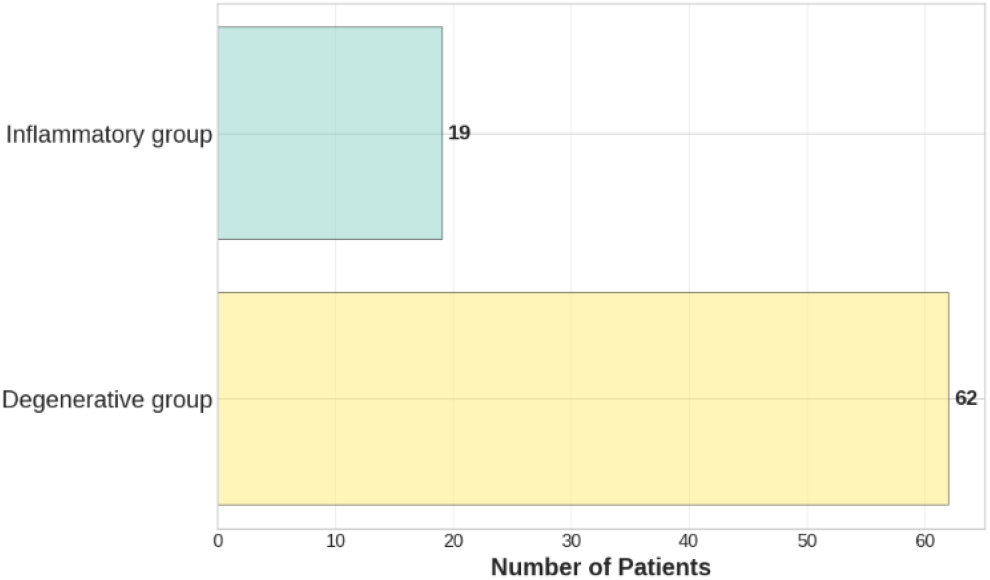
Patients with different conditions were divided into 2 group.

### 2.4 Treatment Protocol

Patients received individualized SynthéXer PEMF treatment sequences selected based on clinical diagnosis. Treatment duration ranged from 1 week to 4 months (Figure 4), with session durations of 15-90 minutes per session. Twelve PEMF treatment sequences were employed. Each sequence was characterized by specific frequency components, waveform parameters, and temporal patterning, with protocols parameters recorded in the device’s treatment log.

**Fig. 4.**
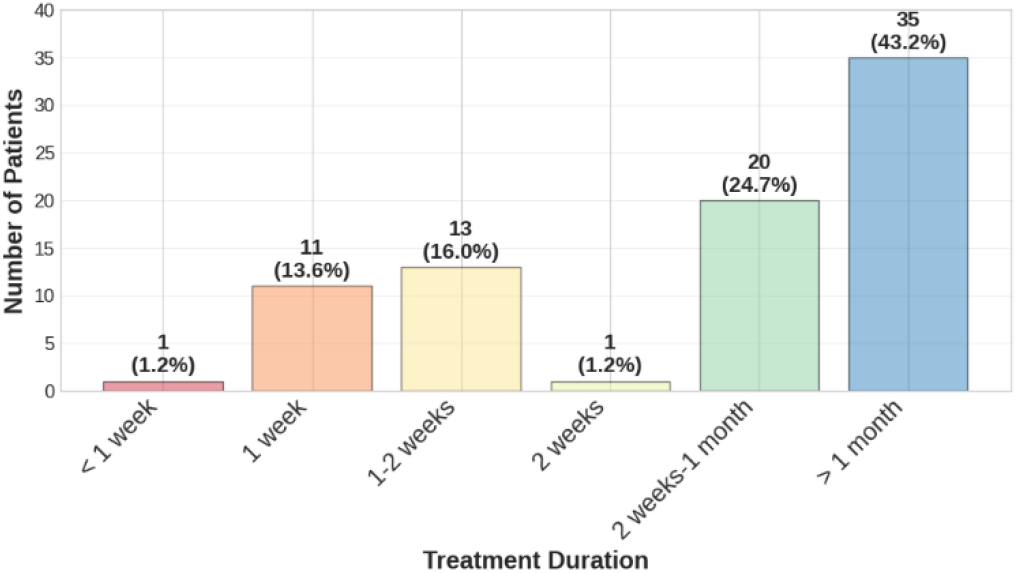
Number of Patients by Treatment Duration: The distribution shows how long the treatment lasted.

### 2.5 Evaluation metrics

Pain intensity was assessed using the Numerical Pain Rating Scale (NPRS), a validated, widely-used self-report measure of pain intensity ranging from 0 (no pain) to 10 (worst imaginable pain) [39,40]. Patients provided NPRS scores immediately prior to initiating treatment (NPRS PRE) and following completion of the treatment course (NPRS POST). For patients reporting pain within a specific range (e.g., 7-8), the higher value was used for conservative statistical analysis (applied to 44/81 subjects).

### 2.6 Statistical Analysis

#### 2.6.1 Descriptive Statistics and Data Characterization

Regarding descriptive statistics, mean, standard deviation, kurtosis, kurtosis SE, skewness, skewness SE and 95% confidence intervals were calculated for NPRS PRE and NPRS POST scores across the full sample and by clinical subgroup (Tab.1 and Tab.7).

**Tab. 1.** Descriptive statistics.

| VARIABLE | MEAN | STD DEV. | KURTOSIS S.E. | KURTOSIS | SKEWNESS S.E. | SKEWNESS |
| --- | --- | --- | --- | --- | --- | --- |
| PRE | 8.07 | ±1.65 | 2.88 | 0.53 | -1.22 | 0.27 |
| POST | 1.79 | ±1.67 | 1.48 | 0.53 | 1.17 | 0.27 |
| Δ NPRS | -6.28 | ±2.09 | 0.45 | 0.53 | 0.57 | 0.27 |
| N. SESSIONS | 15.59 | ±8.53 | -0.75 | 0.53 | 0.39 | 0.27 |
| DURATION | 41.79 | ±35.58 | -0.83 | 0.53 | 0.74 | 0.27 |

**Tab. 2.** Reduction in pain and number of sessions for centers.

| Center | n. | $\Delta \text{NPRS}$ | n. sessions |
| --- | --- | --- | --- |
| Turin | 11 | $-7.18 \pm 1.25$ | $16.91 \pm 9.12$ |
| Arezzo | 14 | $-7.14 \pm 1.23$ | $16.43 \pm 8.85$ |
| Somma Lombardo | 27 | $-6.56 \pm 1.52$ | $20.56 \pm 6.12$ |
| Milazzo | 29 | $-5.10 \pm 1.48$ | $9.52 \pm 5.31$ |

**Tab. 3.**
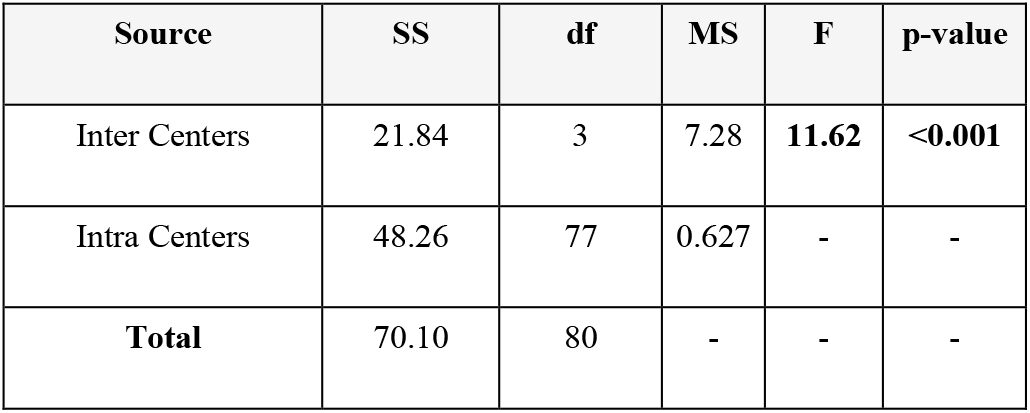
One-Way ANOVA on Δ NPRS.

| Source | SS | df | MS | F | p-value |
| --- | --- | --- | --- | --- | --- |
| Inter Centers | 21.84 | 3 | 7.28 | **11.62** | **<0.001** |
| Intra Centers | 48.26 | 77 | 0.627 | - | - |
| **Total** | **70.10** | **80** | - | - | - |

**Tab. 4.**
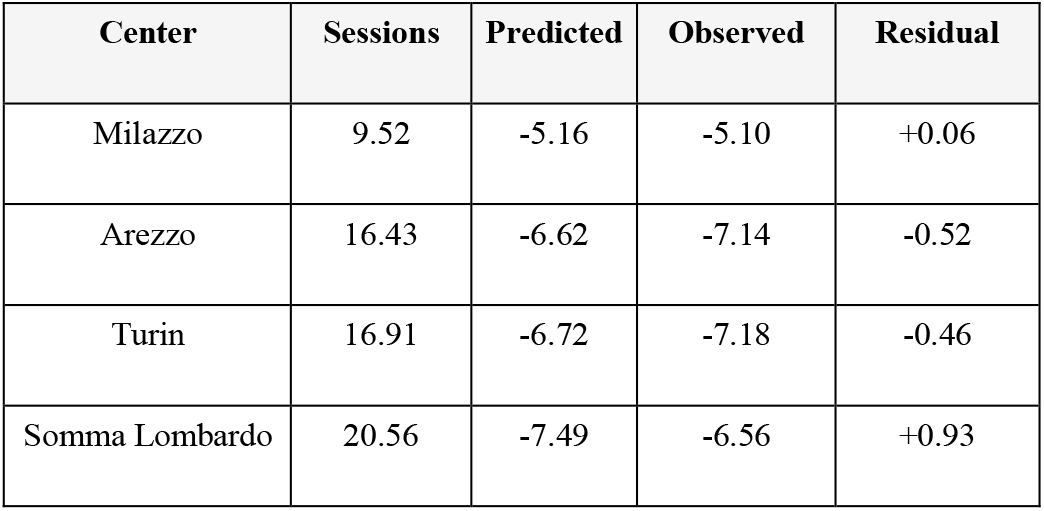
Linear Regression Model.

| Center | Sessions | Predicted | Observed | Residual |
| --- | --- | --- | --- | --- |
| Milazzo | 9.52 | -5.16 | -5.10 | +0.06 |
| Arezzo | 16.43 | -6.62 | -7.14 | -0.52 |
| Turin | 16.91 | -6.72 | -7.18 | -0.46 |
| Somma Lombardo | 20.56 | -7.49 | -6.56 | +0.93 |

**Tab. 5.** ANCOVA: Adjusting for Sessions Frequency.

| Effect | F | p-value | Partial $\eta^2$ |
| --- | --- | --- | --- |
| Sessions (covariate) | 25.78 | <0.001 | 0.380 |
| Center (unadjusted) | 11.62 | <0.001 | 0.312 |
| Center (adjusted) | 5.87 | 0.001 | 0.112 |

**Tab. 6.** Mean and standard deviations of NPRS values before and after treatment and corresponding pain reduction in the full cohort. NPRS: Numerical Pain Rating Scale; Δ: Post-Pre Treatment variation.

| NPRS | Mean | Std Dev | 95% CI |
| --- | --- | --- | --- |
| PRE | 8.07 | $\pm 1.65$ | [7.70-8.44] |
| POST | 1.79 | $\pm 1.67$ | [1.41-2.17] |
| $\Delta \text{ NPRS}$ | -6.28 | $\pm 2.09$ | [-6.77, -5.79] |

**Tab. 7.** Comparison of mean NPRS values and respective standard deviations between the two subsamples under examination (Inflammatory and Degenerative) before and after treatment. NPRS: Numerical Pain Rating Scale; Δ: Post-Pre Treatment variation.

|  | NPRS | Mean | Std Dev |
| --- | --- | --- | --- |
| **Degenerative**<br>(n=62) | PRE | 8.21 | $\pm 1.72$ |
| | POST | 1.68 | $\pm 1.70$ |
| | $\Delta$ NPRS | **-6.53** | **$\pm 2.08$** |
| **Inflammatory**<br>(n=19) | PRE | 7.63 | $\pm 1.34$ |
| | POST | 2.16 | $\pm 1.57$ |
| | $\Delta$ NPRS | **-5.47** | **$\pm 1.97$** |

#### 2.6.2 Inter Center analysis

A statistical analysis was conducted to assess inter-centre heterogeneity in the outcomes of SynthéXer PEMF treatment. First, the equivalence of baseline pain levels between sites was verfied using One-way ANOVA (for normally distributed data), Levene’s Test (for homogeneity of variance) and Kruskal-Wallis test (for non-parametric comparisons). Inter-centre differences in Δ NPRS score were then analysed using One-way ANOVA, followed by Tuckey’s HSD post-hoc tests for pairwise comparisons. To evaluate the influence of treatment frequency, a simple linear regression between the number of sessions and pain reduction was performed. Finally, an analysis of covariance (ANCOVA) was conducted to adjust for the center effect for session frequency, with statistical significance set at p<0.05.

#### 2.6.2 Within-group and Subgroup Analyses

Changes within-group from PRE to POST were assessed with: i) Two-factor ANOVA (between-subjects factor: clinical subgroup; within-subjects factor: time) to evaluate main effects and interactions; ii) Pearson correlation coefficients to examine relationships between treatment duration/session count and magnitude of pain reduction; iii) Cohen’s d effect sizes to quantify the magnitude of clinical response, considering a p-value < 0.05 to be statistically significant.

IBM SPSS Statistics (version 4.1, 1991) was used for all analyses.

## 3. Results

### 3.1 Inter-Center analysis

This multicentric analysis (Figure 5) confirmed statistically significant inter-center heterogeneity in SynthéXer PEMF treatment outcome (F(3,77) = 11.62, p < 0.001). However, this variation can be explained by differences in treatment protocol implementation, specifically the number of therapeutic sessions delivered per patient.

**Fig. 5.**
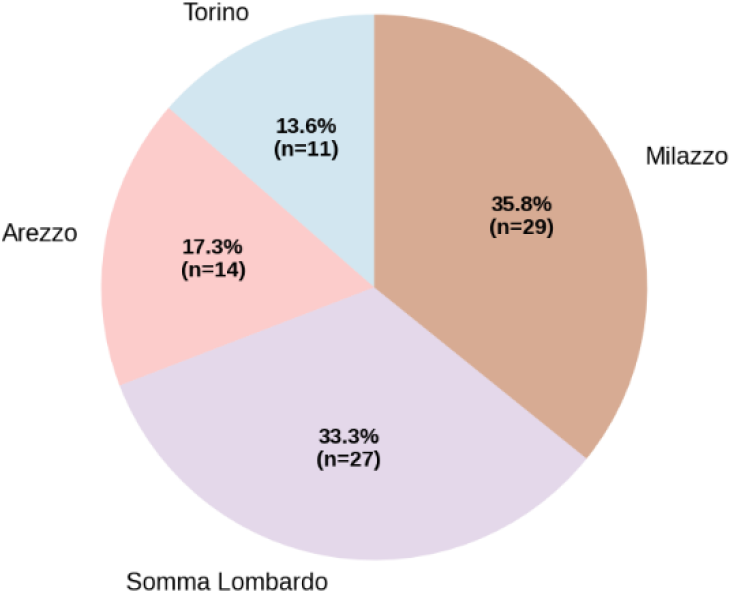
Number of Patients by Medical Center.. Geographic distribution of the four Italian rehabilitation centers participating in the PMS study, with number of enrolled patients per center.

Baseline equivalence testing as One-way ANOVA (F(3,77)=1.33, p-value=0.271), Levene’s Test (F=0.52, p-value=0.668) and Kruskal-Wallis (H=2.21 0 p-value= 0.529) confirmed that initial pain levels were statistically comparable across centers (p > 0.05), excluding patient selection bias as source of outcome differences.

Table 2 summarizes that all centers achieved clinically meaningful pain reduction (71.7%–84.5%) and responder rates between 66% and 93%. However, Milazzo—with the lowest mean number of sessions (9.52 ± 5.31)—exhibited the smallest pain reduction (Δ NPRS = −5.10 ± 1.48), while Somma Lombardo, despite the highest session frequency (20.56 ± 6.12), performs worse than predicted (Table 4), suggesting the need to define the patient mean features to stratify the analysis.

#### 3.1.1 Statistical analysis of between-center heterogeneity

To assess the presence of significant differences among the study groups, a one-way analysis of variance (ANOVA) was conducted. The results, summarized in Table 3, reveal a statistically significant effect of the grouping variable on the measured outcome (F(3, 77) = 11.62, p < 0.001). The between-group variability (Inter Centers) accounted for a substantial portion of the total variance, with a sum of squares (SS) of 21.84 and a mean square (MS) of 7.28. In contrast, the within-group variability (Intra Centers) was lower, with an MS of 0.627. The highly significant p-value indicates that at least one group mean differs significantly from the others. These findings suggest that the grouping factor has a meaningful impact on the observed variable, warranting further investigation through post-hoc analyses to identify specific group differences.

The Tukey’s Honestly Significant Difference (HSD) test is a single-step, multiple comparison procedure that calculates all pairwise differences between group means while controlling the family-wise error rate through the studentized range distribution. Post-Hoc Tukey HSD analysis revealed a significant divergence in outcomes among treatment centers: Milazzo differed statistically from all other centers (p<0.001) while Arezzo, Turin, Somma Lombardo exhibited no significant differences among themselves (p > 0.05).

This pattern aligns with the results of a simple linear regression model (Table 4), where the number of sessions emerged as the primary explanatory variable for changes in pain intensity (Δ NPRS). Specifically, the model Δ NPRS = −3.15 (SE = 0.39, p < 0.001) – 0.211 × Sessions (SE = 0.025, p < 0.001) revealed that each additional treatment session was associated with a 0.21-point greater reduction in pain, with a strong negative correlation (r = −0.674) and an explained variance of 45.4% (R^2^ = 0.454, Adjusted R^2^ = 0.447).

To isolate the contribution of treatment frequency from center-specific effects, an ANCOVA was conducted adjusting for session frequency (Table 5). The results confirmed session frequency as a dominant covariate (F = 25.78, p< 0.001, partial η^2^ = 0.380), explaining approximately 64% of the observed inter-center heterogeneity. While the unadjusted center effect remained significant (F = 11.62, p < 0.001, partial η^2^ = 0.312), adjustment for session frequency attenuated this effect (F = 5.87, p = 0.001, partial η^2^ = 0.112), indicating that residual variability (11%) may reflect additional operational or patient-specific factors that do not compromise data validity.

In conclusion, inter-center heterogeneity in pain reduction outcomes is clinically meaningful and primarly driven by Milazzo’s lower-intensity treatment protocol. Session frequency alone explains 45% of the total outcome variance, underscoring its central role. However, the persistence of a residual center effect (11%) after adjustment suggests that additional factors—such as patient characteristics or local protocol adaptations—may contribute to the observed variability. These findings support the validity of multicentric data pooling when appropriate statistical adjustments for treatment dosage are applied, while highlighting the need for further stratification to account for unmeasured confounders.

### 3.2 Full Cohort analysis (n=81)

Following exclusion criteria application, the analyzed cohort comprised 81 patients with chronic pain due to inflammatory (n=19) or degenerative (n=62) conditions. Treatment intensity varied across the cohort (Figure 4): mean number of treatment sessions was 15.59 ± 8.53, and mean treatment duration was 41.79 ± 35.58 days.

In the analysis that involved full cohort (n=81), the mean NPRS score decreased significantly from 8.07 ± 1.65 (PRE) to 1.79 ± 1.67 (POST), representing a reduction of 6.28 ± 2.09 points (p < 0.001) (Table 6 and Figure 6). The Effect Size was estimated by the Cohen’s d and was 3.1, indicating a very large effect size, substantially exceeding conventional thresholds (small: 0.2; medium: 0.5; large: 0.8).

**Fig. 6.**
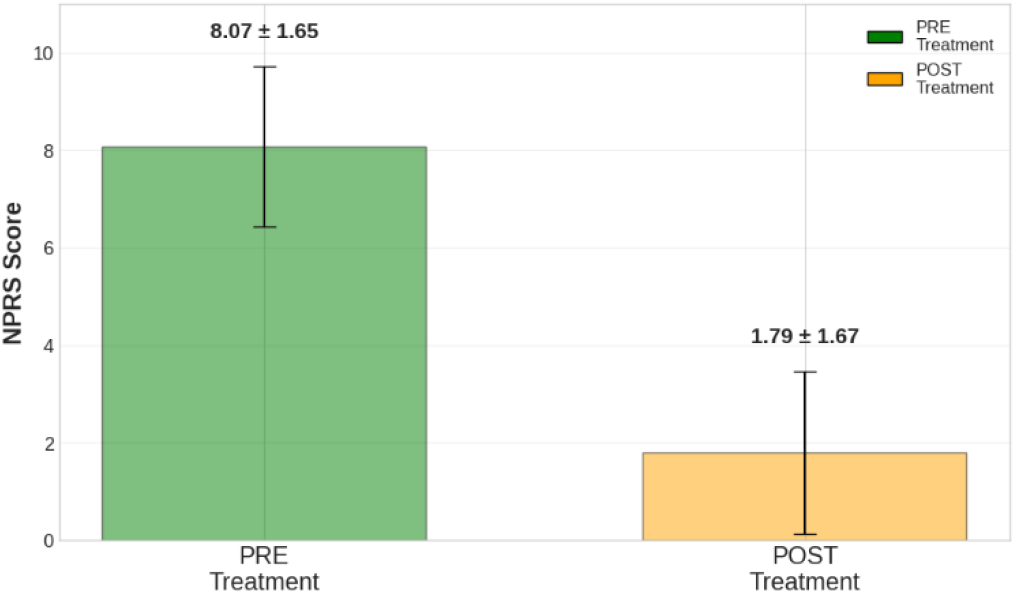
Pain Reduction Across Full Cohort. Mean Numerical Pain Rating Scale (NPRS) scores pre-treatment (PRE) and post-treatment (POST) for the entire patient cohort (n=81). Error bars represent standard deviation. ***p<0.001 by analysis of Variance.

Regarding the Individual-Level Response, 80/81 patients (98.8%) demonstrated pain reduction (Δ NPRS > 0); 79/81 patients (97.6%) achieved clinically meaningful reduction (Δ NPRS ≥ 2); 66/81 patients (81.5%) reached post-treatment NPRS ≤ 3 (mild/no pain).

### 3.3 Subgroup analysis

Both subgroups (Degenerative and Inflammatory) demonstrated substantial pain reductions following treatment (Table 7, Figure 7). To assess potential differences in pain reduction between subgroups, an ANOVA analysis was conducted (see Section 3.4).

**Fig. 7.**
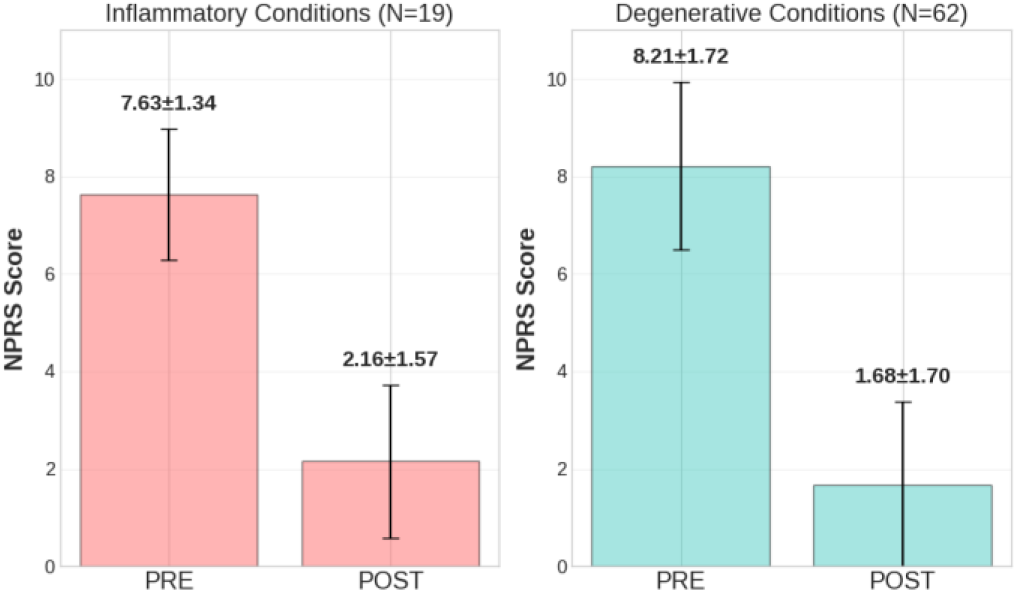
Subgroup analysis of NPRS scores pre- and post-treatment for Inflammatory (n=19) and Degenerative (n=62) conditions. Both groups exhibited substantial pain reduction, with no significant difference between subgroups (p=0.886, ANOVA).

#### 3.3.1 Analysis of Variance (ANOVA)

Table 8 presents the results of a two-factor ANOVA examining the effects of **Time** (PRE vs POST) and **Clinical Subgroup** (Inflammatory vs Degenerative) on pain reduction.

**Tab. 8.** Two-factor Analysis of Variance (ANOVA) results.

| Source | SS | df | MS | F | p |
| --- | --- | --- | --- | --- | --- |
| Time | 1048.15 | 1 | 1048.15 | 498.56 | <0.001* |
| Sub-group | 0.07 | 1 | 0.07 | 0.02 | 0.886 |
| Time × Sub-group | 8.15 | 1 | 8.15 | 3.88 | 0.052 |

The main effect of Time was highly significant (p < 0.001), confirming substantial pain reduction across the cohort. The Time × Subgroup interaction approached statistical significance (p = 0.052), suggesting that the clinical subgroup may modestly influence the trajectory or magnitude of the treatment response, meriting further investigation in future studies.

The degenerative group exhibited slightly greater absolute reductions than Inflammatory group (Table 7). However, this difference was not statistically significant (Table 8, ANOVA subgroup effect p = 0.886).

### 3.5 Correlation Analysis

Relationships between treatment parameters and clinical response were examined to assess potential dose-response effects.

Pearson correlation coefficients were calculated to quantify the strength and direction of these associations.

Statistically significant negative correlations were observed, indicating that a greater number of treatment sessions and a longer overall treatment duration were associated with larger reductions in pain (Table 9). Specifically, the number of sessions correlated with pain reduction (r = −0.358, p = 0.001), as did the total treatment duration (r = −0.312, p = 0.005). This pattern is consistent with a dose-response relationship, supporting the biologically plausible hypothesis that increased PEMF exposure enhances therapeutic efficacy.

**Tab. 9.** Correlation analysis between treatment frequency, treatment duration and pain reduction.

| Variables | Coeff. (r) | p value |
| --- | --- | --- |
| Pain reduction (Δ NPRS) vs Session count | -0.358 | 0.001** |
| Pain reduction (Δ NPRS) vs Treatment duration | -0.312 | 0.005** |

The dose-response relationship is further illustrated in Figure 8, where an exponential interpolation model was applied to visualize the data. Panel (A) demonstrates how pain reduction varies as the number of sessions increases, with a calculated half-life of 6.6 sessions—indicating that approximately 50% of the maximum pain reduction is achieved after this number of sessions. Similarly, Panel (B) shows the relationship between pain reduction and treatment duration, with a half-life of 5.7 days. These findings suggest that both the frequency and cumulative duration of PEMF treatment contribute to clinical improvements, although the effect may plateau after a certain threshold.

**Fig. 8.**
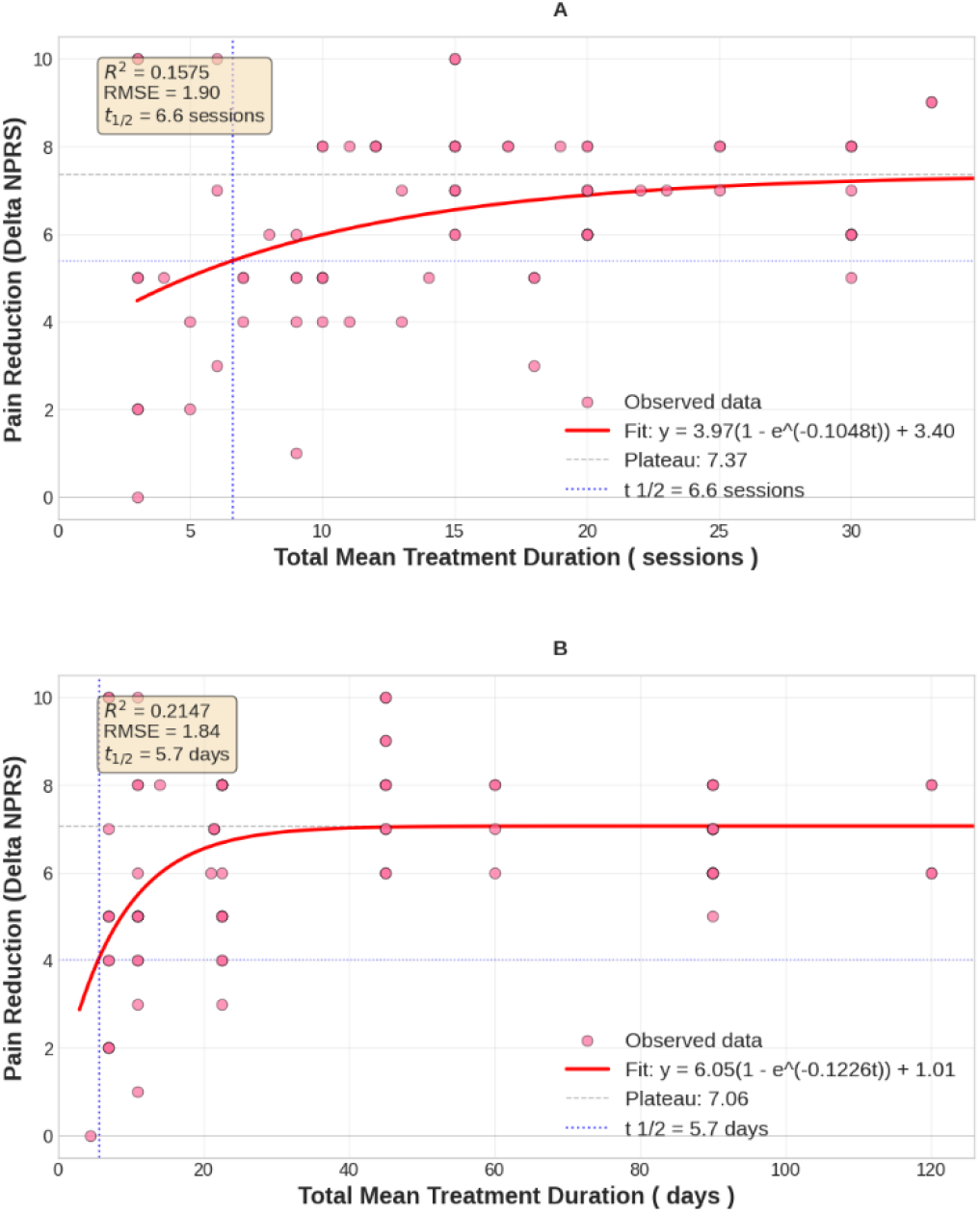
Dose-Response Relationship. The interpolation model used is the Exponential Model. (**A**) The figure shows how Pain Reduction varies as the number of sessions increases. The half-life is equal to 6.6 sessions. (**B**) The figure shows how Pain Reduction varies as the days increase. The half-life is equal to 5.7 days.

### 3.6 Safety Analysis

No adverse events were reported during or following PEMF treatment, and no patient discontinued therapy due to adverse effects. Tolerability was uniformly rated as good across all participating centers, indicating that the treatment was well-received by patients. The absence of adverse events aligns with previous literature on PEMF therapy, which generally reports a favorable safety profile for this modality.

## 4. Proposed Biological Mechanism: Integration of Clinical Data with Molecular and Epigenetic Evidence

The magnitude of pain reduction observed in this PMS cohort exceeds typical responses to many standard analgesic interventions, suggesting the involvement of substantive biological mechanisms. While the observational design precludes definitive causal inference, the consistent response magnitude across heterogeneous patient populations and the emerging evidence for PEMF sequence-specific epigenetic effects in vitro collectively suggest the activation of biological framework meriting consideration.

### 4.1 KDM6B-mediated epigenetic remodeling in response to specific PEMF sequences

A recent in vitro study demonstrated that exposure of promonocytic U937 cells to specific PEMF sequences generated by the SynthéXer system results in a constellation of co-ordinated molecular changes (Figure 9).

**Fig. 9.**
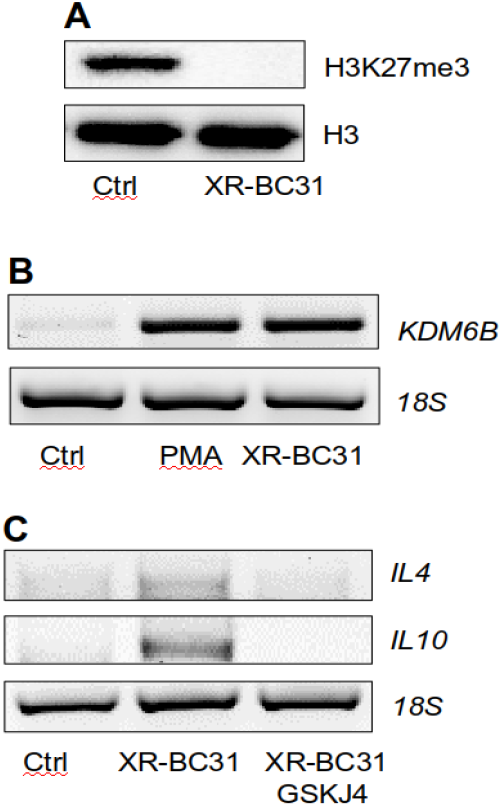
(**A**) Representative Western blot analysis of H3K27me3 and histone H3 in U937 cells exposed 4 days to the XR-BC31 sequence (once a day), by means of the SynthéXer® system. (**B**) Representative RT-PCR analysis of KDM6B expression in U937 cells untreated (Ctrl), treated 48 hours with 10 nM PMA (PMA) or exposed 4 days to the XR-BC31 sequences. (**C**) Representative RTPCR analysis of IL4 and IL10 expression in U937 cells not exposed (Ctrl) or exposed to XRBC31 sequence in the absence or in the presence of the selective KDM6B inhibitor, GSKJ4. 18S was used as housekeeping gene.

#### Histone Demethylase Activation

The H3K27me3-specific demethylase KDM6B is significantly upregulated following PEMF exposure with specific frequency/modulation patterns [37].

#### Chromatin Remodeling

KDM6B-mediated demethylation of H3K27me3 results in a global reduction of this repressive histone mark, opening chromatin structure and facilitating transcription of specific gene sets.

#### Anti-inflammatory Macrophage Differentiation

PEMF exposure promotes differentiation of promonocytic cells toward an anti-inflammatory (M2 macrophage) phenotype, characterized by elevated expression of scavenger receptors, mannose receptors, and reduced pro-inflammatory effector functions.

#### Altered Cytokine Secretion

These epigenetically reprogrammed macrophages exhibit substantially increased secretion of anti-inflammatory interleukins, particularly IL-10 and IL-4, while reducing expression of pro-inflammatory mediators including TNF-α, IL-1β, and IL-6 (Figure 10) [37].

**Fig. 10.**
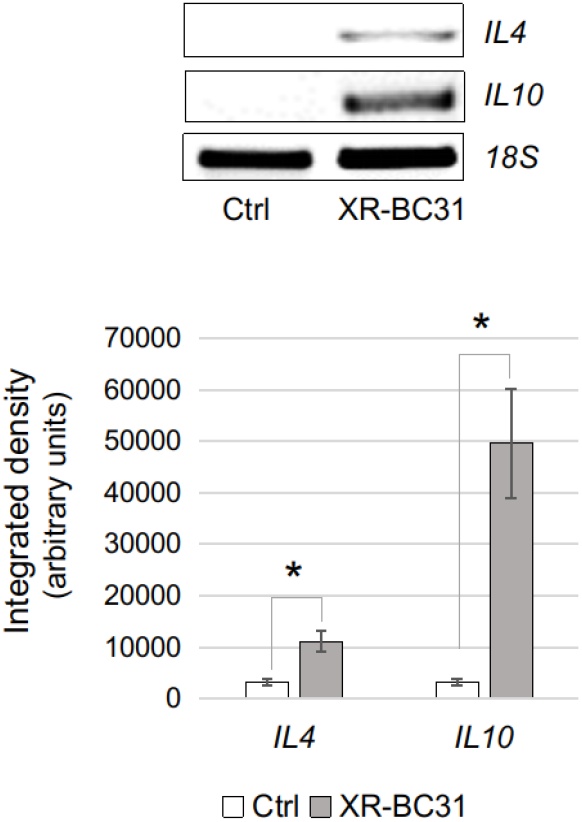
Representative RT-PCR and densitometric analysis of IL4 and IL10 expression (cropped images are parts of the same gel) in control (Ctrl) and XR-BC31 sequence exposed U937 cells (XR-BC31). 18S was used as housekeeping gene. In the graph, each bar represents mean of three independent experiments. * p≤0.05

Importantly, these effects were highly sequence-dependent: identical PEMF exposure protocols using slightly different modulation parameters failed to induce these changes, highlighting the critical importance of specific electromagnetic waveform characteristics [37]. This sequence-specificity argues against non-specific physical effects and implicates structured biological signaling mechanisms.

### 4.2 Proposed Mechanistic Link: from KDM6B to pain resolution

If PEMF exposure via SynthéXer promotes KDM6B-dependent epigenetic reprogramming in tissue macrophages (and possibly other immune cells), one would predict: 1. Enhanced macrophage plasticity and capacity for M2 differentiation. 2. Increased expression of anti-inflammatory cytokines (IL-10, IL-4, IL-13). 3. Reduced expression of pain-promoting pro-inflammatory mediators (TNF-α, IL-1β, IL-6). 4. Restoration of inflammatory homeostasis. 5. Reduction in pain signaling from peripheral tissues.

This mechanistic chain, while requiring direct experimental confirmation in vivo, provides a plausible biological bridge between in vitro epigenetic observations and in vivo clinical pain reduction. The proposed mechanistic molecular pathway aligns with and extends existing literature on PEMF effects.

#### PEMF and Myeloid Cell Function

Numerous studies document PEMF-induced alterations in macrophage and dendritic cell phenotype and cytokine secretion, with generally increased anti-inflammatory responses [18–20, 23, 41–43].

#### Ion Dynamics and Signal Transduction

The well-established models of PEMF-membrane interaction (ion resonance, Lorentz force effects, electrochemical perturbation) [11, 13–17] provide mechanistically plausible pathways through which electromagnetic stimulation could alter intracellular calcium dynamics, ROS production, and signal transduction cascades upstream of epigenetic changes.

#### ROS and Redox-Sensitive Signaling

Recent evidence suggests PEMF-induced modulation of reactive oxygen species (ROS) and activation of redox-sensitive kinases may contribute to downstream transcriptional changes [16, 17, 44–46]. Epigenetic “writers” and “erasers” (including KDM6B) are subject to redox regulation, providing a potential mechanistic link [47].

#### Adenosine Signaling

PEMF-induced changes in extracellular adenosine concentration and adenosine receptor expression have been documented [48], and adenosine signaling is known to modulate macrophage polarization and pain signaling [49]. Collectively, these converging lines of evidence suggest that sequence-specific PEMF exposure may activate a coordinated biological program involving membrane perturbation, signal transduction, epigenetic remodelling of chromatin, modulation of immune cell phenotype and pain resolution.

### 4.3 Clinical Correlates: why Inflammatory and Degenerative conditions show similar responses

A notable finding in our PMS cohort was that both inflammatory and degenerative pain conditions responded robustly to PEMF, with minimal subgroup differences. How does the proposed KDM6B mechanism account for this pattern?

Inflammatory conditions (rheumatoid arthritis, soft tissue inflammation) feature prominent immune cell infiltration and pro-inflammatory cytokine production; thus, direct promotion of macrophage M2 polarization [50] and anti-inflammatory IL-10 production via KDM6B activation would directly address disease pathophysiology.

Degenerative conditions (osteoarthritis, discopathies) feature substantial low-grade inflammation, including macrophage infiltration of degenerated tissues and elevation of pain-producing mediators [36, 51, 52]. Furthermore, degenerative joint disease is increasingly understood to involve altered macrophage phenotype and dysregulated inflammatory homeostasis [53]. Thus, epigenetic promotion of anti-inflammatory macrophage differentiation would also benefit degenerative pain conditions.

The near-equivalence of treatment effects across inflammatory and degenerative subgroups thus supports a mechanistic model centered on the activation of macrophage cells and antiinflammatory effectors, rather than acting on mechanisms specific to a single disease category. Indeed, in inflammatory conditions the pain is more immune-mediated contributing to central sensitization, whereas in degenerative conditions the pain is more mechanical and structural in origin due to cartilage breakdown and bone-on-bone contact. Nevertheless, PEMF treatment efficiently suppresses pain discomfort in both diseases groups.

### 4.4 Limitations of the Proposed Framework and Requirements for Future Investigation

Several critical gaps between current evidence and mechanistic confirmation must be acknowledged.

#### Direct In Vivo Validation required

The proposed mechanism is derived from in vitro molecular studies and in vivo clinical observations. Direct demonstration that PEMF exposure promotes KDM6B upregulation and H3K27 demethylation in macrophages of treated patients would strengthen causal inference. This might be accomplished via immune cell isolation from peripheral blood or synovial fluid, with assessment of KDM6B expression, H3K27me3 status, and macrophage polarization markers.

#### Sequence-Specificity in Vivo

While in vitro studies demonstrate sequence-dependent effects, it remains to be demonstrated whether the specific SynthéXer sequences used clinically (e.g., XR-BC31) are indeed responsible for KDM6B activation in vivo. Comparative studies with different PEMF protocols would be informative.

#### Contribution of Other Epigenetic Mechanisms

KDM6B is one of many epigenetic regulators. PEMF might also affect DNA methylation (via regulation of specific enzymes), other histone modifiers, and non-coding RNA expression. Comprehensive epigenomic profiling of PEMF-exposed cells would clarify the full landscape of epigenetic changes.

#### Central vs. Peripheral Mechanisms

While the proposed framework emphasizes peripheral tissue macrophage effects, PEMF exposure might also influence central nervous system immune cells (microglia, perivascular macrophages), central sensitization, and descending pain modulation. Mechanistic studies should not assume purely peripheral action.

Although promising, current evidence derives from observational clinical data and in vitro studies. Randomized controlled trials with sham-PEMF controls, blinded outcome assessment, and mechanistic biomarkers would be required to definitively establish efficacy and mechanism.

## 5. Discussion

### 5.1 Clinical Significance of observed pain reduction

The mean pain reduction of 6.28 ± 2.09 NPRS points represents a substantial clinical improvement. To contextualize: current literature suggests that a reduction of ≥2 points on the NPRS is considered clinically meaningful [54], and reductions of 4-5 points are considered very large [55]. Our cohort achieved substantially greater reductions, with 97.6% of patients demonstrating ≥2-point reductions and 81.5% achieving mild or absent pain post-treatment (NPRS ≤ 3).

Importantly, these results were obtained in a real-world, heterogeneous population across multiple centers, supporting generalizability. No adverse events were reported, suggesting a favorable safety profile compared to many pharmacological alternatives. The negative correlations between treatment duration/session count and pain reduction magnitude (r = −0.31 to −0.36, p < 0.005) suggest a dose-response relationship: longer treatment courses and more frequent sessions were associated with greater pain reductions. The exponential interpolation model showed how the frequency and cumulative duration of PEMF treatment contribute to clinical improvements, although the effect may plateau after a certain threshold.

The study involved patients with diverse etiologies of chronic pain: osteoarthritis, rheumatoid arthritis, soft tissue inflammation, vertebral pathology, neuropathic pain, and others. Despite this heterogeneity, responses were remarkably consistent, with substantial pain reduction across subgroups. This pattern differs from many pharmacological interventions, which typically show variable efficacy across etiologically distinct conditions. The ability to produce consistent analgesic responses across such diverse pain phenotypes suggests action on a common biological pathway rather than disorder-specific mechanisms.

### 5.3 Observational Study Design: Strengths and Limitations

This PMS study provides valuable real-world evidence of PEMF effectiveness but, as an observational study without control group, cannot definitively establish causation or quantify placebo effects.

The study’s strengths include its multi-center design, which enhances the generalizability of the findings, and the use of a standardized assessment tool (NPRS) consistently applied across all sites. A large effect size was observed, alongside a consistent response across diverse pathologies, underscoring the robustness of the results. Systematic safety monitoring and correlations between treatment parameters and clinical outcomes further support a dose-response relationship.

This study has several limitations. The absence of a control group or sham-PEMF arm limits the ability to attribute observed effects solely to the intervention. Additionally, the lack of blinding for both patients and outcome assessors introduces potential bias. Pain measurements relied on self-report, which is susceptible to expectancy and recall bias. The study also lacked mechanistic biomarkers, such as immune phenotyping or epigenetic assessments, which could provide deeper insights into the underlying biological mechanisms. Finally, the heterogeneous patient population and variability in treatment protocols may obscure potential subgroup effects.

### 5.4 Integration with Mechanistic Framework

While the observational clinical data do not prove mechanism, they provide motivation for proposing testable biological hypotheses. It is worth noting that the patients cohort received PEMF treatments using the same device (SynthéXer) and PEMF sequences similar to those used to treat in vitro U937 cells. This provided a basis for proposing that a comparable mechanism may be triggered in patients as well. The proposed epigenetic framework—centered on sequence-specific PEMF promotion of KDM6B activity, H3K27 demethylation, anti-inflammatory macrophage differentiation, and altered cytokine production—offers a coherent explanation for the observed clinical responses and is grounded in peer-reviewed cellular and molecular evidence. Critically, this framework makes specific, testable predictions: A) PEMF exposure with SynthéXer-specific sequences should promote KDM6B expression in immuno cells in vivo. B) Treated patients should show increased circulating or tissue anti-inflammatory macrophages. C) Treatment-responsive patients should show reduced pro-inflammatory cytokine production. D)These biomarker changes should correlate with degree of pain reduction. Future studies designed to test these predictions would provide stronger evidence for mechanism while simultaneously validating the clinical efficacy suggested by this PMS data. The clinical findings from this PMS study—namely, the large and consistent pain reduction across diverse etiologies, the dose–response relationship, and the absence of serious adverse events—are not only clinically meaningful but also biologically coherent in light of the KDM6B-centered epigenetic model outlined in Section 4.

Specifically, the observed transdiagnostic efficacy aligns with a mechanism that targets a shared pathophysiological node: dysregulated tissue inflammation mediated by macrophage polarization. Whether the primary driver is autoimmune (e.g., rheumatoid arthritis), mechanical-degenerative (e.g., osteoarthritis), or neuropathic, persistent pain in these conditions is frequently sustained by pro-inflammatory cytokine signaling (TNF-α, IL-1β, IL-6) and impaired resolution pathways. The proposed PEMF-induced upregulation of KDM6B, leading to H3K27me3 demethylation and M2 macrophage skewing, offers a unified explanation for the broad therapeutic response.

### 5.5 Relationship to Existing PEMF Literature

The proposed KDM6B-centered mechanism complements and extends existing literature. Decades of preclinical and clinical research document PEMF effects on bone healing, soft tissue repair, and pain reduction [7–9, 56–60]. However, most prior investigations were conducted with empirically optimized but incompletely characterized electromagnetic waveforms, limiting mechanistic understanding.

The current work emphasizes the critical importance of electromagnetic waveform parameters in determining biological response. This sequence-specificity paradigm—demonstrated convincingly in recent molecular study [37]—suggests that prior heterogeneous PEMF research results may partially reflect use of suboptimal waveforms. Optimized sequences might produce superior clinical responses than earlier PEMF systems.

The proposed centrality of KDM6B connects PEMF effects to contemporary understanding of immune cell epigenetics. KDM6B has recently emerged as a therapeutic target in cancer immunology, rheumatoid arthritis research, and resolution of inflammation [31–35, 50, 61]. The hypothesis that PEMF activates KDM6B-dependent immune remodeling reprogramming thus aligns with cutting-edge immunological science.

### 5.6 Clinical and Translational Implications

Even though the proposed mechanism is substantiated, several important clinical implications emerge.

#### Optimization of PEMF Protocols

Identifying the specific electromagnetic sequences most effective in promoting KDM6B activation and M2 macrophage differentiation could guide development of next-generation PEMF systems with enhanced efficacy.

#### Mechanistic Biomarker Development

Circulating or tissue epigenetic markers (KDM6B expression, H3K27me3 status, macrophage phenotyping) could be developed as biomarkers of PEMF response, potentially enabling prediction of individual treatment responsiveness and optimization of protocols.

#### Synergistic Therapeutic Approaches

Understanding PEMF action on macrophage immune homeostasis could motivate combination approaches, e.g., PEMF combined with dietary anti-inflammatory agents, anti-inflammatory pharmaceuticals, or other immunomodulatory therapies.

#### Broader Application

Assuming that KDM6B activation is indeed central to PEMF benefits, this mechanism might extend to other pain conditions and potentially to inflammatory diseases beyond pain, such as autoimmune and inflammatory bowel disorders (for which macrophage polarization is pathogenic).

### 5.7 Questions and Future Directions

Despite the promising findings, several critical questions remain unanswered. From a mechanistic perspective, it is essential to determine whether PEMF-treated patients exhibit increased circulating anti-inflammatory macrophages and elevated IL-10 expression. Additionally, it is unclear whether epigenetic markers in immune cells, such as KDM6B and H3K27me3, correlate with clinical response, providing a molecular basis for the observed effects.

In vivo confirmation of these mechanisms is equally important. Does PEMF exposure promote KDM6B-dependent demethylation in tissue macrophages in vivo, and can this be demonstrated in accessible tissues, such as peripheral blood or skin biopsies, or even through non-invasive molecular imaging? Addressing these questions would bridge the gap between in vitro observations and clinical relevance.

Another key area for future research is the optimization of PEMF sequences. Which specific sequences are most effective in promoting KDM6B activity, and can rational design principles be applied to generate superior sequences? Answering this could significantly enhance the therapeutic potential of PEMF.

The potential effects of PEMF on the central nervous system also warrant investigation. Does PEMF influence central immune cells, such as microglia, or modulate central pain processing and descending pain inhibitory pathways? Exploring these aspects could reveal novel mechanisms underlying PEMF’s analgesic effects.

Finally, to move from observational evidence to definitive proof of efficacy and mechanism, randomized controlled trials with sham-PEMF controls, blinded assessments, and mechanistic biomarkers are urgently needed. Such studies are required to move from observational evidence to definitive proof of efficacy and mechanism.

## 6. Conclusions

This multi-center PMS study confirmed that pulsed electromagnetic field therapy with the SynthéXer system produces pain reduction in patients with chronic pain due to inflammatory and degenerative conditions. The magnitude of clinical response (6.28 ± 2.09 NPRS point reduction; Cohen’s d = 3.1) is notable and consistent across a heterogeneous patient population.

While the observational study design precludes definitive causal attribution, the convergence of clinical data with emerging cellular evidence of sequence-dependent epigenetic effects suggests a plausible biological framework. Specifically, we propose that sequence-specific PEMF exposure promotes KDM6B-mediated histone demethylation and chromatin remodeling in immune cells, facilitating anti-inflammatory macrophage differentiation and altered expression of regulatory cytokines (IL-10, IL-4). This mechanism, operating at the intersection of biophysical stimulus, epigenetic remodeling, and immune system function, could explain the pronounced and consistent pain reduction observed.

However, substantial additional investigation is necessary to validate and extend this framework. Direct demonstration of KDM6B activation and epigenetic changes in PEMF-treated patients, confirmation of immune cell phenotypic changes, and definitive linkage of these biomarkers to clinical pain reduction would substantially strengthen causal inference. Randomized controlled trials with sham-PEMF controls and comprehensive mechanistic assessment are required to establish definitive efficacy and elucidate biological action.

Ultimately, these findings suggest that PEMF therapy represents a promising non-pharmacological, non-invasive approach to chronic pain management, with emerging evidence for a coherent biological mechanism. Further investigation to confirm and refine mechanistic understanding will likely enhance clinical efficacy and guide development of optimized next-generation systems.

### 6.1. Data availability

The raw data of the results presented in the paper are available upon request to the corresponding author.

### 6.2. Funding

This research did not receive any specific grant from funding agencies in the public, commercial, or not-for-profit sectors.

### 6.4. Author contributions

Material preparation and data collection were performed by Ethidea Srl. Analysis was performed by Ethidea Srl. The first draft of the manuscript was written by CS, and both authors commented on previous versions of the manuscript. AF provided supervision and review.

## Data Availability

All data produced in the present study are available upon reasonable request to the authors. The dataset analyzed in this preprint derives from MDR post-market surveillance (PMS) activities and was provided to the manufacturer in fully anonymized form. The analysis is retrospective and non-interventional, based on routine use, with pre- and post-treatment pain measures; no additional procedures were introduced for research purposes. The manufacturer did not access personal data, and no re-identification key was available. Therefore, ethics committee approval and informed consent were not required/not applicable for this anonymized PMS dataset.

## 7. Acknowledgements

We thank Dott. Massimo Balma and Dott. Gianfranco Fonte for helpful discussion and comments.

